# Essential newborn care in Sidama, Ethiopia: Findings from a community-based cross-sectional household survey

**DOI:** 10.64898/2026.02.06.26345520

**Authors:** Hirut Gemeda, Yemisrach Shiferaw, Andargachew Kassa, Yaliso Yaya, Achamyelesh Gebretsadik

## Abstract

**Abstract:** *Objective:* To describe the existing status and associated factors influencing the utilization of four WHO-recommended essential newborn care among mothers of infants aged 45 days to one year in rural Sidama, Ethiopia.

*Design:* A community-based cross-sectional household survey was conducted in June and July 2023. Data were collected through interviewing mothers of infants using pretested questionnaire. Participants were selected through a multi-stage sampling. Data were analyzed using Stata V.15.

*Setting:* Selected rural kebeles of Bilate Zuriya, Boricha, Hawassa Zuriya and Shebedino districts of Sidama, Ethiopia.

*Participants:* 1,821 mothers of infants aged 45 days to one year.

*Primary outcome measures:* the proportion of babies who received some or all components of the four WHO-recommended essential newborn cares.

**Results:** Of 1,821 mothers, 53.9% (981/1,821) reported that their newborns had received immediate and thorough drying, 52.7% (959/1,821) indicated immediate skin-to-skin contact, and 46.9% (854/1,821) revealed that babies initiated breastfeeding within one hour after birth. However, only 2.3% (42/1,821) of mothers reported delayed cord clamping. No newborn received all four. Only 15% (273/1,821) reportedly received at least three of the four, 38.8% (706/1,821) received two, 33.2% (605/1,821) received one, and 13% (237/1,821) have not received any of the care. Facility delivery (RRR=5.26; 95% CI: 1.11, 8.89), Proximity to a facility (RRR=1.70; 95% CI: 1.09, 2.67), living in communities with higher wealth (RRR=11.74; 95% CI: 3.09, 44.63), insurance coverage (RRR=6.54; 95% CI: 2.25, 19.06), and education levels (RRR=7.75; 95% CI: 2.57, 23.34) were significantly associated with utilization of three.

**Conclusion:** The utilization rate of essential newborn care in rural Sidama is unacceptably low. Individual and community level factors were significantly associated with the use. A comprehensive strategy must therefore address the identified factors.

**Strengths and limitations of the study:** This study has methodological strengths including the uses, a strong community-based study design with a large sample size (N=1,821) and complete response rate, rigorous data quality assurance through electronic collection (Kobo tool box). Further we carefully selected and trained data collectors to minimize social desirability bias, and included a number of variables relevant for policy considerations, and used of multilevel modeling to account for hierarchical data structure. However, the researchers acknowledged several limitations and implemented strategies to alleviate them. Reliance on maternal self-report introduces potential recall and social desirability biases, which were addressed through specific interviewer techniques and by focusing on memorable events. Moreover, the lack of direct observational cross-validation for clinical practices such delayed cord clamping remains a constraint.

## Introduction

The World Health Organization (WHO) defines Essential Newborn Care (ENC) as a set of simple, evidence-based interventions provided at birth and during the immediate postnatal period to reduce preventable newborn morbidity and mortality [1, 2]. These interventions comprise four important elements of care: immediate, thorough drying and stimulation of a baby to prevent hypothermia; delayed cord cutting to enhance neonatal blood volume and iron stores; early initiation of breastfeeding within the first hour of life; and thermal care through skin-to-skin contact to maintain temperature and physiological stability. Together, these essential care practices address the major risks faced by newborns in the first hours and days after birth and serve as important indicators of the quality of newborn care [2-4]

Immediate and thorough drying and stimulation at birth is critical for preventing hypothermia, particularly in resource-limited settings where access to advanced resuscitation services is often critically limited [5] Delayed cord cutting, defined as clamping the umbilical cord one to three minutes after birth, allows continued placental transfusion of blood to the newborn [4]. This practice has been shown to increase neonatal hemoglobin levels and iron stores and to reduce the risk of anemia in infancy, which is particularly important in settings where childhood anemia is highly prevalent [6, 7]. Early initiation of breastfeeding provides colostrum, which is rich in antibodies and essential nutrients, supports immune development, and has been associated with reduced neonatal morbidity and mortality [8, 9]. Thermal care, especially skin-to-skin contact between the mother and newborn, helps regulate body temperature, stabilize physiological functions, and strengthen early mother–infant bonding [10].

This WHO guidance and other related national and international efforts have contributed to significant improvements in child survival worldwide over the past two decades [11, 12]. Despite the gains, mortality and severe morbidity around childbirth remain unacceptably high, particularly in low-income and middle-income countries (LMICs), including those in sub-Saharan Africa. Recent estimates indicate that more than 80% of neonatal deaths occur in LMICs, with Africa accounting for approximately 43% of global neonatal deaths [13]. Nearly half of all deaths among children under five years of age occur during the neonatal period, with the highest risk concentrated in the first 24 hours after birth. Most neonatal deaths are caused by preventable or treatable conditions such as complications of prematurity, birth asphyxia, and severe infections [14, 15].

This burden in low-income settings highlights a critical gap between global progress in child survival and the effective delivery of life-saving interventions at the time of birth, where it matters most. Evidence consistently shows that simple, low-cost interventions delivered at or immediately after birth can substantially improve newborn health and survival [16, 17]. Nevertheless, access to and use of these interventions remain limited in many rural and underserved settings, where health systems often face constraints of resource shortages and poor utilization of existing services.

In Ethiopia, the challenges related to poor access to essential care and underutilization remain substantial, especially in rural areas such as Sidama. A study conducted in 2020 reported that 56% of births took place at home without skilled birth attendance, and among newborns delivered at home, only 2.4% received essential newborn care [18]. These findings highlight important gaps in the delivery of immediate newborn care at the community level.

In Sidama, essential newborn health care is particularly concerning because the majority of the population resides in rural areas and access to quality health services is limited. A study conducted in 2018 reported a neonatal mortality rate of 41 per 1,000 live births in Sidama [19], indicating a still high burden of preventable neonatal deaths in the region.

Several previous studies conducted in Ethiopia have examined essential newborn care, with a primary focus on parents’ knowledge, awareness, and attitudes towards ENC practices [20, 21]. While these findings made a valuable contribution to understanding demand-side intentions, there remains limited evidence on the proportion of newborns who actually receive the recommended essential care practices. Moreover, few studies have systematically examined how individual, household, neighborhood (community), and broader contextual factors, such as residence in particular villages and districts, influence ENC utilization [22, 23]. We aim to describe the existing status of the use of the four WHO-recommended essential newborn care practices and factors influencing their use in rural areas of Sidama Regional State, Ethiopia.

## Methods

### Study Setting

This study was conducted in rural communities of Sidama Regional State, about 400 kilometers south of Ethiopia’s capital, Addis Ababa. The Sidama Region is home to approximately 4.6 million residents as of 2023, with Hawassa serving as its regional capital. The area comprises 37 districts, seven town administrations, and 602 *kebeles* (the smallest administrative unit). The region has a diverse population, with many residents living in rural and remote areas who primarily depend on agriculture and face challenges due to limited access to quality healthcare services [24]. Its healthcare infrastructure includes 21 district hospitals, 135 health centers, and 551 health posts. Health posts are staffed by two to four health extension workers (HEWs). The survey was conducted across four purposively selected districts (Woredas in the Ethiopian context) in the region (See Fig. 1). The districts are Bilate Zuriya, Boricha, Hawasa Zuriya, and Shebedino.

**Fig. 1.**
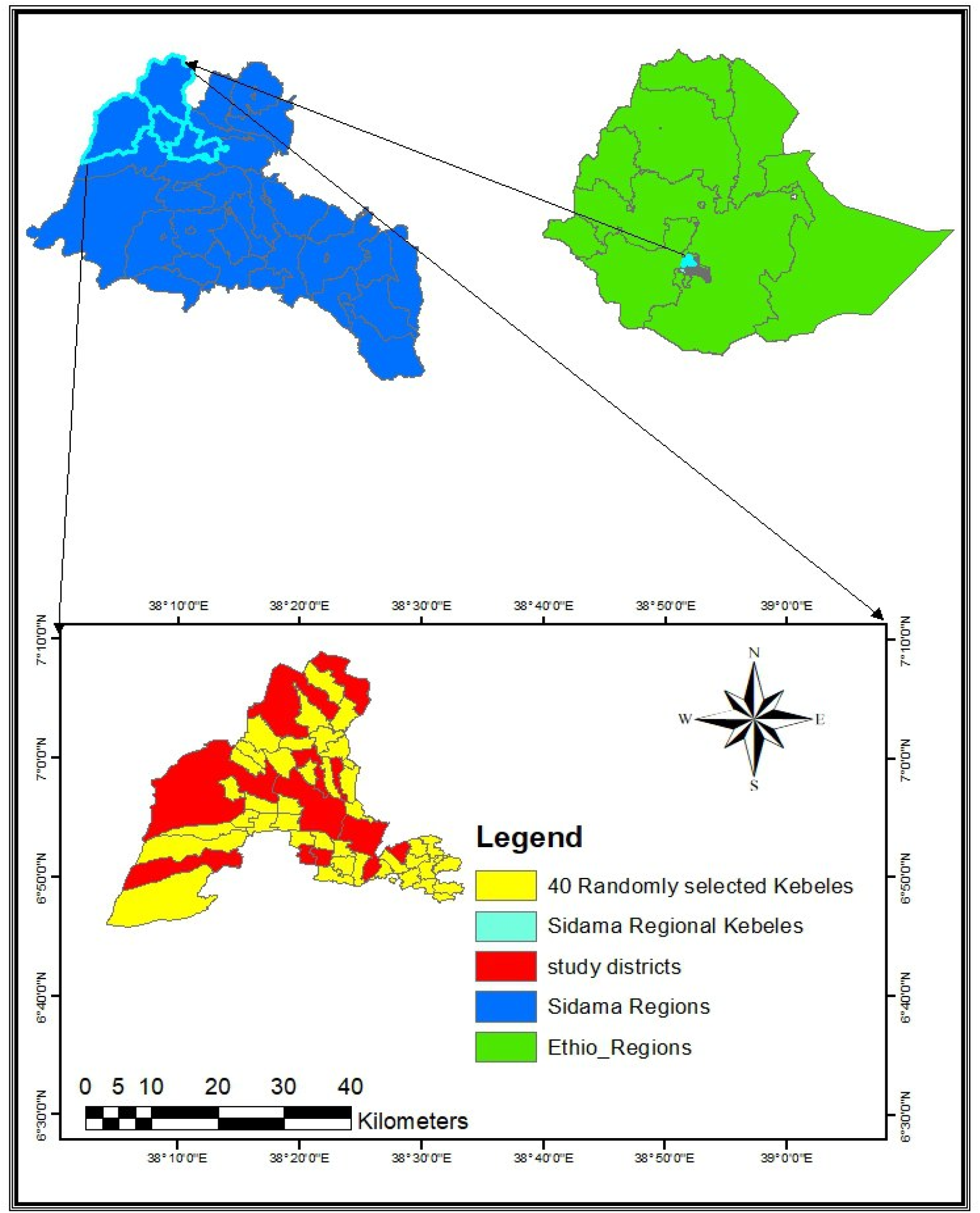
The map showing the research area in Ethiopia’s Sidama regional state. The authors prepared this figure and have granted permission to share it under a CC BY 4.0 license. Source: Shapefile obtained from the Sidama Regional Health Bureau in 2022.

#### Study Period

The household survey was conducted from June 4th to July 16th, 2023.

#### Study Design

We conducted a community-based cross-sectional household survey.

#### Source populations and target population

Eligible mothers or caregivers and their newborn pairs residing in the selected four districts in the Sidama Regional State. We interviewed respondents who had a live-born infant aged 45 days to one year at the time of data collection.

#### Sample size determination and sampling procedure

##### Sample size determination

The sample size was calculated using the formula for estimating a single population proportion in OpenEpi, based on an assumed prevalence of 28% [22] for the essential newborn care practice, a 95% confidence level. With a design effect of 2.0, a target sample of 1,721 respondents was determined. However, since this study was part of a larger research project, with a sample size of 1821 respondents, we included in this study the entire sample of a cluster-randomized trial to assess the effectiveness of a set of community-based interventions on maternal and newborn healthcare [24].

##### Sampling techniques and participant selection

A multistage cluster sampling design was employed to obtain a representative sample of mother-infant pairs. The sampling process consisted of four stages. First, four districts (Woredas)— Bilate Zuriya, Boricha, Hawassa Zuriya, and Shebedino were purposively selected from the Sidama Region because they represented typical Sidama rural areas demographically and in terms of infrastructure. Second, *kebeles* were randomly selected from each district using a simple random sampling technique, facilitated by computer-generated random numbers in SPSS as follows. (See Table 1)

**Table 1.** Sampling techniques and participant selection.

| Woreda (districts) | No.of *Kebeles* | No.of participant mothers |
| --- | --- | --- |
| Bilate Zuriya | 8 | 381 |
| Boricha | 6 | 287 |
| Hawassa Zuriya | 12 | 576 |
| Shebedino | 14 | 577 |
| Total | 40 | 1821 |

##### Inclusion criteria

We included a respondent mother or, if the mother was a permanent resident of the selected *kebele*, had a live birth within 45 days to one year before the interview, and was able to complete the interview.

##### Exclusion criteria

The study applied the following exclusion criteria: non-permanent residents or those living in the selected districts for less than six months; mothers with newborns aged of less than 45 days, and mothers who experienced a stillbirth in their most recent pregnancy.

#### Variables

##### Exposure variables

Exposure variables included based on their previous association with essential newborn care in other studies. These are newborn sex, socio demographic characteristics of the mothers such as; maternal age, education, occupation, wealth index, walking distance to a health facility, health insurance, village literacy ratio and obstetric characteristics are gravidity, reported gestational age, Antenatal care (ANC), place of delivery, Postnatal care (PNC) [25-27].

Gravidity: the number of times a woman has been pregnant, regardless of whether the pregnancies were interrupted or resulted in a live birth.

Gestational age (GA) at birth: duration of pregnancy completed in weeks when the baby was born.

Antenatal care (ANC): is a health service provided to pregnant women in the continuum of maternity care

Postnatal care (PNC): is a care offered to a woman and her infant during the postnatal period.

The health extension workers (HEWs): are women with a high school education who have undergone a one-year training in general health promotion and disease prevention. They work according to the national implementation manual to optimize the health extension Program, which mainly emphasizes health promotion, disease prevention, and increasing maternal and child healthcare utilization [28]

##### Outcome Variables

Immediately and thoroughly drying (ITD): is defined as whether the baby will be completely wiped with a cloth prior to placental delivery.

Immediate skin-to-skin contact (ISSC): placing of the newborn on the mother’s chest and/or abdomen before delivery of the placenta.

Early initiation of breastfeeding (EIBF): is defined as the infant initiating breastfeeding within the first hour postpartum

Delayed cord clamping (DCC): cord clamping performed approximately 1–3 minutes after birth [4]

##### Measurement of variables used for multilevel-multinomial logistic regression

The outcome variable is measured by the proportion of babies who received some or all components of the four WHO-recommended essential newborn care. 1/ immediately and thoroughly drying, 2/skin-to-skin contact, 3/early initiation of breastfeeding, and 4/delayed cord clamping [1, 2].The investigator measured the outcome as follows: For each component of essential newborn care, we assigned a score of 1 if the newborn received that care component and 0 if not. To measure overall utilization of essential elements of newborn care, a composite score was calculated for each newborn based on the number of essential care elements they received. This variable was coded from 0 to 4, where a score of ‘0’ indicated that the newborn received none of the elements, a ‘1’ indicated receipt of one, a ‘2’ indicated two, a ‘3’ indicated three, and a ‘4’ signified that the newborn received all four essential elements of care.

##### Data Collection Instruments and Procedures

Data were collected via face-to-face interviews at the respondents’ homes using a structured questionnaire. Data were collected using the Kobo Collect survey application (version 2021.2.4; The KoBo Toolbox Consortium, 2021) [29]. The questionnaire was developed in the English language based on a review of relevant literature and adapted to the local context. The questionnaire was then translated into the local language, “*Sidamo affoo,”* and back-translated to English to ensure accuracy.

Data collection was carried out by trained diploma-holder enumerators selected from the local community under the supervision of health professionals. The data collectors received two days of training that covered the study’s objectives, the practical and ethical aspects of conducting interviews, the clarity of interview questions, and methods to reduce socially desirable responding where survey respondents give answers, they believe will be viewed favorably rather than their true thoughts or behaviors [30].

#### Data structure and analysis

The data had a hierarchical structure, with analysis based on clustered individual-level data: individuals were nested within *kebeles* and *kebeles* nested within districts. As such, we conducted a Multilevel Multinomial Regression analysis using a Generalized Structural Equation Model (GSEM). The outcome variable, essential newborn care (ENC) utilization, was a four-category nominal variable indicating a composite variable of the number of ENC a newborn received: newborns who did not receive any care element (ENC =0), received one (ENC = 1), received two (ENC = 2), received three (ENC = 3), and received all four elements (ENC = 4). The outcome was treated as nominal, and the category that received none of the ENC elements (ENC = 0) was specified as the reference category (base outcome). We excluded receiving only one ENC because it is not practically significant and is included only in descriptive reporting. In other words, we compared the groups that received two or three essential care elements with those that received none. Since no group received all four essential care elements, one group (ENC = 4) was excluded from the analysis.

#### Data processing and analysis

Data were exported from the Kobo Toolbox to IBM SPSS Statistics, Version 26 (IBM Corp, 2019) and Stata, Version 15 (Stata Corp, College Station, TX, USA). Following data cleaning and coding in SPSS, descriptive statistics in the form of frequencies and percentages. Initially, binary logistic regression was employed, and any predictor variable with p < 0.25 was retained as a candidate. These candidate variables were subsequently entered into a multivariable logistic regression model to identify factors associated with the outcome.

We fitted multilevel multinomial logistic regression models using generalized structural equation modelling (GSEM) in Stata. Multinomial logit (mlogit) models were specified with ENC = 0 as the reference outcome. Fixed effects were included for individual-level variables, including sex of infant, maternal age, education, occupation, wealth index, gestational age, antenatal care (ANC), place of delivery, walking distance to the nearest health facility and community-level covariates such as wealth index, health insurance, health extension workers ratio, educational status, walking distance to the nearest health center.

To account for clustering in the data, random intercepts were specified at multiple levels. Model building proceeded sequentially. First, a fixed-effects only model was fitted using individual and community-level variables only. Second, a model including the first model and a *kebele*-level random intercept was estimated. Third, a model including the second model and a district-level random intercept was considered. Finally, a fourth and more complex model including random intercepts at both district and *kebele* within district levels was fitted. *Kebele* within district identifiers were constructed to ensure correct nesting.

#### Model comparison and decision

We assessed model fitness to the data by using Akaike’s Information Criterion (AIC) and Bayesian Information Criterion (BIC). Lower values of AIC and BIC indicated better model fit. We selected the model with the lowest AIC/BIC ratio as the best fit model to our data. (Supplementary Table 1).

#### Assessment of clustering

The intra-class correlation coefficient (ICC) was calculated using the latent-variable approach, assuming an individual-level variance of π^2^/3. ICCs were computed from the variance to quantify the proportion of unexplained variance attributable to district-level and village-level clustering. All analyses were conducted using Stata, Version 15 (Stata Corp, College Station, TX, USA), and statistical significance was assessed at the 5% level, with Relative Risk Ratios (RRRs) and 95% confidence intervals (CIs).

#### Quality Assurance

To ensure the reliability and validity of the questionnaire, a small pilot study was conducted in another *kebele* with characteristics similar to those of the study sites. The questionnaire was refined based on the results of the pilot study’s pretest. Data quality was further assured through daily checks for completeness and consistency by the supervisory team.

## Result

### Socio demographic characteristics

The study included 1821 participant mothers with a newborn aged 45 day to one year at the time of data collection. Among the newborns, a slight majority were male 1046 (57.4%). The majority of mothers, 667 (36.6%), were between 25 and 29 years of age 963 (52.9%) had a formal education, and 1480 (81.3%) had no formal paid employment during the time of the survey.

### Obstetric characteristics

The majority of respondents, 1317 (72.3%), had a history of pregnancy experience more than once. Among these, only 322 (17.7%) of mothers had received antenatal care four or more times during the pregnancy, and 1192 (65.5%) delivered in a health facility. A quarter of mothers, however, 458 (25.2%), received postnatal care from health extension workers, and two-thirds, 1089 (59.8%), lived in households located more than 30 minutes of walking distance from a health center, (See Table 2).

**Table 2.** Individual characteristics of mother-infant pairs of the selected rural districts of Sidama region, Ethiopia in 2023 (N=1821)

| Variables | Frequency | % |
| --- | --- | --- |
| Sex of infant |  |  |
| Female | 775 | 42.6 |
| Male | 1046 | 57.4 |
| Maternal Age |  |  |
| 15-19 | 41 | 2.3 |
| 20-24 | 395 | 21.7 |
| 25-29 | 667 | 36.6 |
| 30-34 | 486 | 26.7 |
| ≥ 35 | 232 | 12.7 |
| Education |  |  |
| No formal education | 858 | 47.1 |
| Formal education | 963 | 52.9 |
| Occupation |  |  |
| Not employed | 1480 | 81.3 |
| Employed | 341 | 18.7 |
| Wealth index |  |  |
| Poor | 580 | 31.8 |
| Medium | 633 | 34.8 |
| Wealthy | 608 | 33.4 |
| Gravida |  |  |
| Primigravida | 504 | 27.7 |
| Multi-gravida | 1317 | 72.3 |
| Gestational Age |  |  |
| Early | 55 | 3.0 |
| Normal | 1203 | 66.1 |
| Late | 563 | 30.9 |
| Antenatal care (ANC) |  |  |
| < 4 visits | 1499 | 82.3 |
| ≥ 4 visits | 322 | 17.7 |
| Place of Delivery |  |  |
| Home | 629 | 34.5 |
| Health Facility | 1192 | 65.5 |
| Postnatal care (PNC) received<br>from a health extension worker |  |  |
| No | 1363 | 74.8 |
| Yes | 458 | 25.2 |
| Walking distance to the nearest<br>health facility |  |  |
| > 30mins | 1089 | 59.8 |
| ≤ 30mins | 732 | 40.2 |

### Newborns who received each of the essential newborn care elements

The receipt of four essential elements of newborn care was assessed among 1,821 infants. About 53.9% (981/1821) newborns received immediate and thorough drying, 52.7% (959/1821) of newborns obtained immediate skin-to-skin contact with the mother, and 46.9% (854/1821) of newborns started breast-feeding within one hour following birth. Yet only 2.3% (42/1821) of newborn had their umbilical cords clamped within a recommended delay of one to three minutes (See Fig.2).

**Fig. 2.**
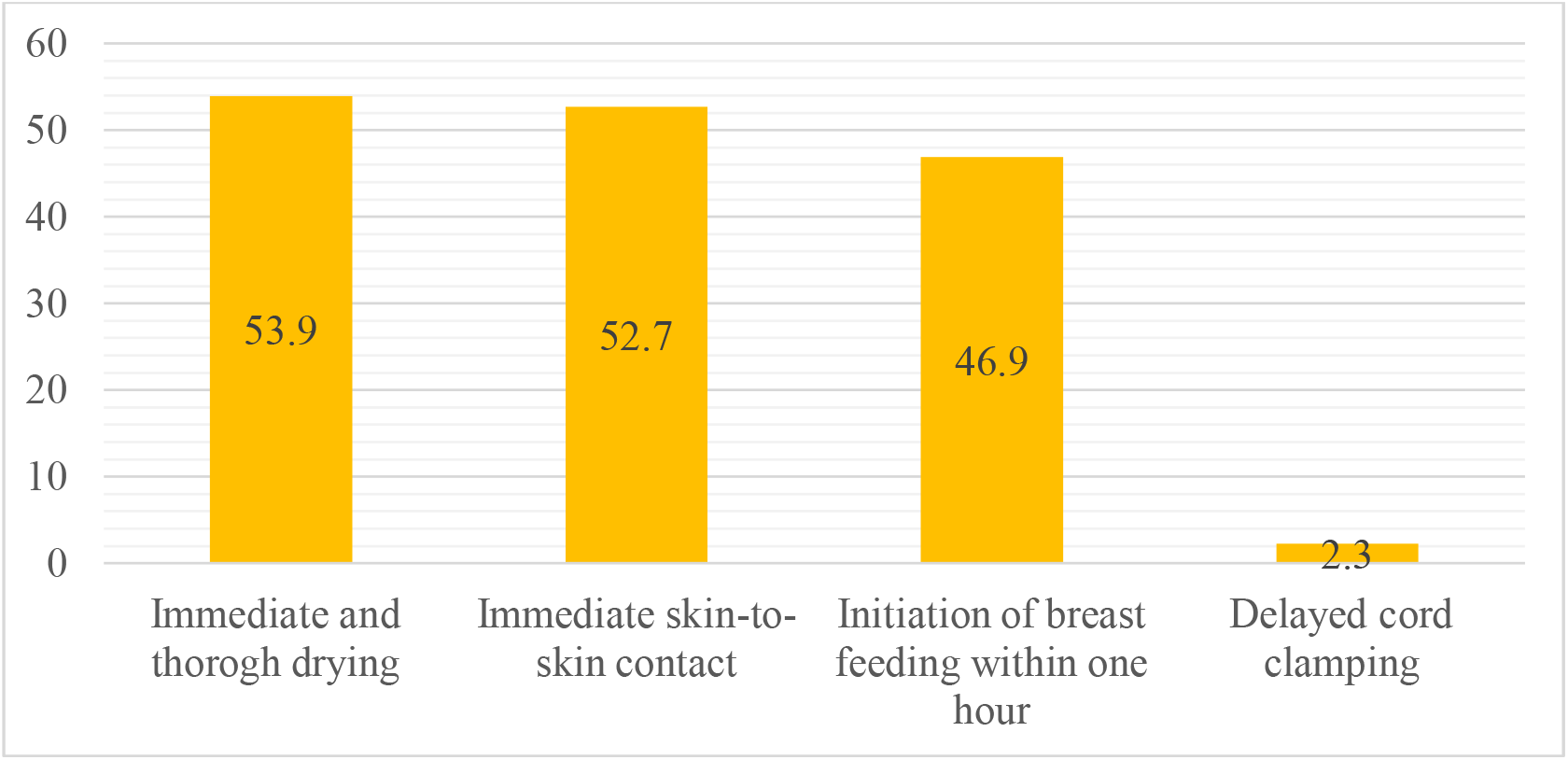
The proportion of newborns who received each of WHO-recommended essential newborn care elements or not in selected Sidama region, Ethiopia in 2023.

### The composite outcome of receiving several elements of ENC

The analysis included 1,821 individuals from 40 villages across four districts. Overall, 13% (237/1821) of mothers reported that their babies received none of the four ENC elements, 33.2% (607/1821) received only one, 38.8% (705/1821) reported two, and 15% (272/1821) three. None of the mothers reported their newborn baby received all four ENC (See Fig. 3).

**Fig. 3.**
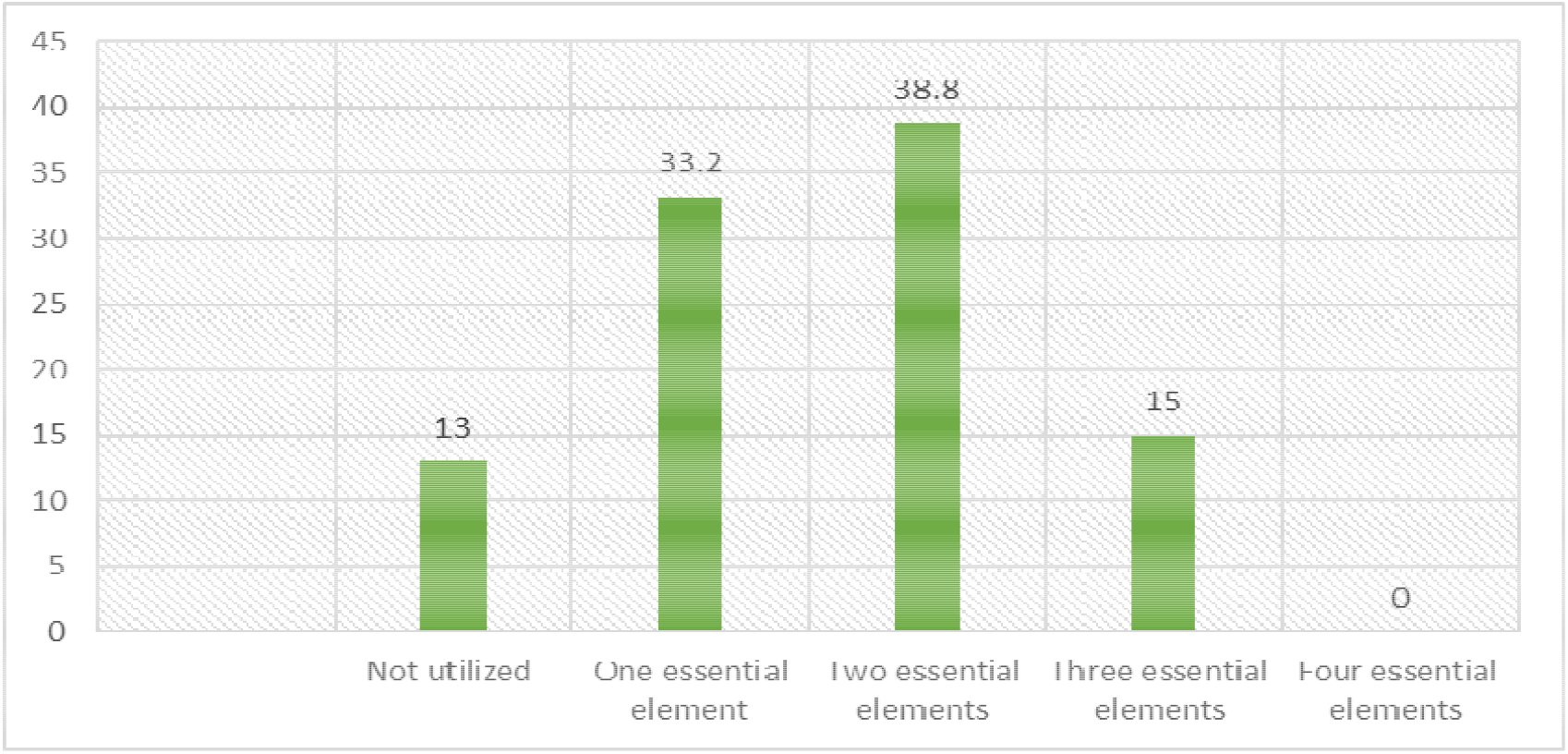
The proportion of newborns who received a set of WHO-recommended essential newborn care elements in Sidama districts, Ethiopia, 2023.

### Factors associated with utilization of a number of essential newborn care (ENC)

Table 2. presents the relative risk ratios (RRRs) and 95% confidence intervals from the final multilevel multinomial logistic regression model. Compared with individuals reporting none users (ENC=0), those reported the use of two or three ENCs gave birth in a facility and resided within less than half an hour walking distance to a health institution, and being in the higher category of household wealth index. At the community level, higher literacy ratio at the village level, village health insurance utilization ratio was significantly associated with the likelihood of newborn babies receiving two or three ENC (See Table 3).

### Random effects and intra-class correlation

Significant clustering of the outcome was observed at both district and village levels. The estimated variance of the district-level random intercept was variance was 4.29, corresponded to an ICC of 0.16 indicating that approximately 16% of the unexplained variation in receiving a number of ENC elements was attributable to between-district differences. The variance of the village-within-district random intercept variance was 3.29, corresponding to an ICC of 0.33 indicating 33% of variability explained by village (Kebele) difference. Compared with the fixed-effects–only model, inclusion of a village-level random intercept resulted in an improvement in model fit, as indicated by a lower ration of AIC/BIC (supplementary Table 1).

**Table 3.** Multilevel Multinomial Regression on WHO-Recommended essential newborn care (ENC) Utilization and associated factors.

| Individual and community level factors | Use two ENC RRR (95% CI) | Use three ENC RRR (95% CI) |
| --- | --- | --- |
| Sex of newborn |  |  |
| Female | Ref | Ref |
| Male | 1.20 (0.86, 1.68) | 1.33 (0.88, 2.02) |
| Maternal-age |  |  |
| 15-19 | Ref | Ref |
| 20-24 | 1.85(0.71, 4.84) | 1.90 (0.53,6.89) |
| 25-29 | 2.28 (0.89,5.86) | 1.63 (0.46,5.78) |
| 30-34 | 1.61(0.62,4.16) | 1.28 (0.36,4.60) |
| ≥ 35 | 1.74 (0.64,4.70) | 1.28 (0.33,4.87) |
| Maternal Education |  |  |
| No formal education | Ref | Ref |
| Formal education | 0.93 (0.66,1.30) | 1.00 (0.65,1.52) |
| Maternal occupation |  |  |
| Not employed | Ref | Ref |
| Employed | **0.51 (0.35, 0.76) \*\*** | **0.46 (0.27, 0.78) \*\*** |
| Wealth index |  |  |
| Poor | Ref | Ref |
| Medium | 0.84 (0.55,1.29) | 0.72 (0.42,1.22) |
| Rich | 0.68 (0.43,1.08) | 0.70 (0.40,1.23) |
| Reported Gestational Age |  |  |
| Early | Ref | Ref |
| Normal | 0.64 (0.12,3.44) | 0.55 (0.09,3.30) |
| Late | **0.15 (0.03, 0.80) \*** | **0.04 (0.01, 0.28) \*\*** |
| Antenatal care (ANC) |  |  |
| < 4 visits | Ref | Ref |
| ≥ 4 Visits | 1.74 (0.94,3.20) | 1.85 (0.93,3.71) |
| Place of delivery |  |  |
| Home | Ref | Ref |
| Health Facility | **2.68 (1.82,3.95) \*\*\*** | **5.26 (3.11, 8.89) \*\*\*** |
| Distance |  |  |
| >30mins | Ref | Ref |
| ≤30 mins | **2.10 (1.45, 3.03) \*\*\*** | **1.70 (1.09,2.67) \*\*** |
| CLWI |  |  |
| Poor | Ref | Ref |
| Rich | **5.56 (2.20,14.05) \*\*\*** | **11.74 (3.09,44.63) \*\*\*** |
| CBHI |  |  |
| Lower proportion of users | Ref | Ref |
| Higher proportion of users | **4.24 (1.93,9.31) \*\*\*** | **6.54 (2.25,19.06) \*\*** |
| CHEWR |  |  |
| Below the standard | Ref | Ref |
| To the standard | 0.69 (0.35,1.34) | 0.43 (0.17,1.10) |
| CLES |  |  |
| Lower proportion of educated | Ref | Ref |
| Higher proportion of educated | **3.13 (1.39,7.06) \*\*** | **7.75 (2.57, 23.34) \*\*\*** |
| Distance |  |  |
| > 30 minutes | Ref | Ref |
| ≤ 30 minutes | 0.90 (0.44,1.82) | 1.25 (0.48,3.24) |

Note: \*Statistically significant at p value \*p<0.05, \*\*p<0.01, \*\*\*P<0.001
RRR: Relative Risk Ratio, EENC: essential elements of newborn care; WDNHC: walking distance to the nearest health center;
CLWI: community level wealth index; CBHI: community- based health insurance; CHEWR: community level health extension workers ratio; CLES: community level educational status.

## Discussion

In this study, we assessed the status of utilization of the four WHO-recommended essential elements of newborn care following a livebirth and both individual-level and community-level factors associated with the utilization of the care. The findings revealed that nearly half of all newborns received immediate and thorough drying and were placed in immediate skin-to-skin contact with their mother. Conversely, the practice of delayed cord clamping was exceptionally uncommon. Closer to a half of newborns were reportedly initiated of early breastfeeding. No newborn received all four essential care elements. The coverage was fragmented: just over one-third of newborns received two elements of ENC, while a similar proportion received only one element. Only a tenth of babies received at least three components of ENC, and one in eight newborns received none of the four essential newborn care components.

Key factors associated with the utilization include a range of individual and community-level factors. Maternal employment and post-term gestation emerged as strong negative predictors across all levels of care. In contrast, key positive enablers included individual health-seeking behaviors such as institutional delivery and geographic proximity to a health facility. At the community level, high community-based health insurance coverage in the village of residence, and a higher level of village literacy ratio demonstrated positive associations with the utilization of two or more elements of ENC.

As no other evidence has been identified regarding the overall utilization of ENC using composite measurement, we rely primarily on a single metric for the comparative analysis. In this finding, nearly four in ten respondents reported that their newborns received two essential elements of newborn care. A similar estimate of nearly half was also reported in a study conducted in Nepal [2]. Nearly one in six of neonates received three of the four essential elements of newborn care. This finding is comparable to a study conducted in the Nepal, where the rate was one in nine [2]. We also found that a third of neonates used only a single essential element of newborn care, which is lower than the nearly half reported from Nepal in 2017 [2]. The socioeconomic disparities between the study areas could explain this discrepancy. Additionally, our study revealed that a significant minority of neonates did not receive any of the essential elements of newborn care, a situation almost similar to that reported from Nepal where nearly three in hundred [2]. This small difference might be due to variations in policy and the commitment of community health workers or volunteers in Nepal, who actively assist women during and after the birth process.

Furthermore, none of the neonates in our study received all four essential elements of newborn care, a result still nearly similar to the Nepal study, where the rate was just under one in hundred [2]. A possible reason for this could be the insufficient resources in both countries, which may hinder the standard of neonatal care. This includes a lack of medical facilities, supplies, and skilled healthcare workers.

In the current findings, health facility delivery is the powerful predictor of utilization of essential elements of newborn care, a result consistent with previous reports from across sub-Saharan Africa and Asia [31, 32]. The relationship we observed where facility delivery had greater effect on the utilization of two (RRR= 2.68) and three (RRR= 5.26) essential elements compared to the use underscores the importance of skilled birth attendance as the cornerstone of improving ENC utilization and hence newborn outcomes[4]. This shows that delivering in a facility allows trained staff to provide multiple recommended care practices together.

This study found that, distance was a significant barrier. People who resided far away from a health facility were less likely to use essential newborn care services. Existing evidence supports this finding[33]. In this study the finding revealed that there is paradoxical reverse association between mothers being employed in a paid job and utilization essential elements of newborn care of the newborn babies. This contradicts research findings often linking higher socioeconomic status such as employment with better health outcomes [34]. In the study area context, this may indicate that employed mothers are likely engaged in low-wage, labor-based employment that could present structural barriers, such as a lack of maternity leave, time constraints, and the economic necessity to return to work quickly postpartum. This fact may also indicate that the type of employment may have limited their ability to seek antenatal care and deliver in a facility, suggesting that economic engagement without supportive social policies does not automatically translate into better health-seeking behaviors [35].

Our study also revealed that there was a strong positive association between high community-based health insurance (CBHI) coverage and high literacy ratio at the village of residence and ENC utilization. It provides practical support for the role of financial risk protection mechanisms in removing cost barriers, a factor known to obstruct health service utilization in Ethiopia and similar settings [36, 37]. This suggests that policies aimed at expanding CBHI enrollment could be a highly effective strategy for improving newborn health outcomes.

### Strengths and limitations of the study

This study has methodological strengths including the uses, a strong community-based study design with a large sample size (N=1,821) and complete response rate, rigorous data quality assurance through electronic collection (Kobo tool box). Further we carefully selected and trained data collectors to minimize social desirability bias, and included a number of variables relevant for policy considerations, and used of multilevel modeling to account for hierarchical data structure. However, the researchers acknowledged several limitations and implemented strategies to alleviate them. Reliance on maternal self-report introduces potential recall and social desirability biases, which were addressed through specific interviewer techniques and by focusing on memorable events. Moreover, the lack of direct observational cross-validation for clinical practices such delayed cord clamping remains a constraint.

### Conclusion and recommendations

The coverage of WHO-recommended essential newborn care in rural Sidama is low. A number of individuals, household, neighborhood, and contextual factors were associated with the utilization of essential newborn care in rural Sidama. We recommend conduct a pilot study within health facilities to identify specific gaps, strengthening the quality of maternal and newborn care qualities such as skilled delivery and essential newborn care practices at health facilities, and improving mechanism of service access to families residing in distant villages from health facilities.

## Supporting information

IRB Certificate of 2022

Survey Questionnair 2023 English version

## Data Availability

All data produced in the present study are available upon reasonable request to the authors

## Acknowledgment

We want to extend our deepest gratitude to the Government of Norway for funding this research through the NORHED SENUPH-II programme, the Sidama Regional Health Bureau, Bilate Zuriya, Boricha, Hawassa Zuriya and Shebedino districts for their facilitating the research and data collectors and study participants for their involvement.

## Ethical Considerations

Ethical approval for the study was obtained from the Institutional Review Board of Hawassa University (Ref. No. IRB/088/14; Date: 07/11/2022). Written informed consent was obtained from literate participants, while oral consent was received from illiterate participants after the study information was read to them.

## Consent for publication

Not applicable

## Availability of data and materials

The data supporting this study can be obtained by contacting the corresponding author with a reasonable request.

## Author Contributions

HG, YS, AK, AG, YY Conceptualization, methodology, and Validation. HG writes the drafting of the manuscript and AG, AK and YY reviewed and edited the manuscript. All authors read and approved the final manuscript.

