## Supplementary Table 1 for "Essential newborn care in Sidama, Ethiopia: Findings from a community-based cross-sectional household survey"

Supplementary Table 1. Model comparison of findings

| Model | Fixed effects | Random effects | AIC | BIC | AIC/BIC |
| --- | --- | --- | --- | --- | --- |
| M0 | Individual + community | None | 4239.923 | 4570.352 | 0.9277 |
| M1 | Individual + community | Village | 4110.964 | 4457.879 | 0.9222 |
| M2 | Individual + community | District | 4162.667 | 4509.617 | 0.9231 |
| M3 | Individual + community | Village+ District | 4103.032 | 4466.467 | 0.9186 |
