## Supplementary material for "Essential newborn care in Sidama, Ethiopia: Findings from a community-based cross-sectional household survey": IRB Certificate of 2022

ሀዋሳ ዩኒቨርሲቲ
ህክምናና ጤና ሳይንስ ኮሌጅ
የምርምር ስነ-ምግባር ገምጋሚ ቦርድ

### HAWASSA UNIVERSITY
College of Medicine and Health Sciences
Institutional Review Board

Ref. No: IRB/088/14
Date: 07/11/2022

Name of Researcher(s): **Achamyelesh Gebretsadik (Ph.D., Assoc. Prof.), Yaliso Yaya (Ph.D., Assoc. Prof.), Yemisrach Shiferaw (MPH), Hirut Gemeda (MSc.)**

Topic of Proposal: ***Cluster randomized trial to improve maternal and new born health through capacity building interventions to community health workers and other resources in Sidama regional state, Ethiopia***

Dear researcher(s),
The Institutional Review Board (IRB) at the College of Medicine and Health Sciences of Hawassa University has reviewed the aforementioned research protocol with special emphasis on the following points:

| | | | | |
|---|---|---|---|---|
| 1. Are all principles considered? | | | | |
| 1.1. Respect for persons: | Yes | ☑ | No | ☐ |
| 1.2. Beneficence: | Yes | ☑ | No | ☐ |
| 1.3. Justice: | Yes | ☑ | No | ☐ |
| 2. Are the objectives of the study ethically achievable? | Yes | ☑ | No | ☐ |
| 3. Are the proposed research methods ethically sound? | Yes | ☑ | No | ☐ |

Based on the aforementioned ethical assessment, the IRB has:

| | | |
|---|---|---|
| **A.** Approved the proposal for implementation | ☑ | -Approval period from **Nov.7/ 2022** to **Nov. 6 /2023** |
| B. Conditionally Approved | ☐ | -Element Approved: **Protocol Version No. 1** |
| **C.** Not Approved | ☐ | -Follow up report expected in 6 **months** |

Obligation of the PI:

1. Should comply with the standard international and national scientific and ethical guidelines
2. All amendment and changes made in protocol and consent form needs IRB approval
3. The PI should report SAE within 3 days of the event
4. End of study, including manuscript should be reported to the IRB

Yours faithfully,

Dawit Jember (Asst. Prof.)
Institutional Review Board Chairperson.

Institutional Review Board
Hawassa University College of Medicine & Health Sciences

☎: + 046 8209290 Website: Fax: + 046 2208755 ✉ 1560 CMHS, Hawassa-Ethiopia

CamScanner
