## Supplementary material for "Essential newborn care in Sidama, Ethiopia: Findings from a community-based cross-sectional household survey": Survey Questionnair 2023 English version

Annex IV: Study Information sheet

**DissertationTitle:** Cluster randomized controlled trial to assess the effectiveness of a package of community-based intervention on newborn healthcare in sidama, ethiopia: the SiMaNeH trial.

Greetings! My name is _________________. I am working as a data collector of PhD student, HirutGemeda who is learning at Hawassa University, College of Medicine and Health science, School of public health. Her main supervisor is:

- Dr. Achamyelesh G/Tsadik, Hawassa University, Ethiopia

**The Objective of the study:** To determine the effect of a package of community-based intervention improve newborn health care practice in Sidama region in 2023-2025.

**Procedure:** The study involves interviewer-administered questionnaire with the data collector that will ask you a set of questions using a structured questionnaire. After signing the consent form, the Data collector will then ask you the relevant questions and your responses will be written on the questionnaire. The interview will take about 30 minutes.

**The Risks and Benefit of the study:** There is no any risk or discomfort that you will face by participating in this research except dedication of time for responding. Any personal information registered will be not be transferred to other bodies and kept confidential. Though there is no direct benefit from this research project, the finding of this study will used to implement intervention and reveal out gap regarding community-based neonatal health care.

**Follow- up time:** The neonate will be followed until the end of 28^th^ day. During the postnatal period, newly delivered neonates will be visited five times. These schedules are: at 1st, 3rd, b/n 7^th^ and 14^th^ and 28^th^ day of birth.

**Privacy, anonymity and confidentiality:** Your name will be kept confidential. The study forms will be kept for five years in the project office in locked cabinet. Only research staff will be able to see those forms. Mrs. HirutGemeda is the principal investigator of this research. If you have any question about the research, you may call Mrs. Hirut at +251-0910274402. If you feel that you been treated unfair or have been hurt by joining the study you may call Dr. Achamyelesh using +251-0911303128 , DrAndargachew at +251-0911338895, and Dr. Yaliso at +(47) 96988845.

**The Rights of participants:** completely free to take part or not in this study. If you decide that you do not want to be part of the study, you are welcome. You are also free to withdraw from the study at any time if you feel that you cannot proceed. Even if you do not want to join the study, you will receive the same quality of medical care from governmental health facilities.

**Compensation:** you will not be paid for your participation this study**.** If you agree to our proposal of enrolling your neonate in our study, please indicate that by putting your signature or your left thumbprint at the specified space below. Have you agreed to participate in the research?

1- No (say thank you)	2- Yes (take informed consent)

Annex V: Informed consent

The objective, benefits, harms, procedures and confidentially of the study has been read and explained to me in the language I comprehend. I further understand that, taking part in this study and withdraw from participating in any time without having reason is purely voluntary. I agree to participate in this study.

Participant:

_______________________________________________	__________________

Signature or left thumbprint of participant	Date

________________________________________________	___________________

Signature of the interviewer (Data collector)	Date

Thank you for your cooperation

The signed copies must be given 1) to the PI and 2) to participants

**In this house hold, did you have a child whose age less than one year old? 0. No	1. Yes**

**If the answer is No, go to the next eligible**

**Annex VII: Questionnaires**

**Section – 1**

Household characteristics

House ID [____/____]

Neonate ID [__/___]

District Name

Kebele (Name)

Sub-kebele(gote/gasha ) code [___,____]

Interviewer name ___________________________ signature..........................................

Supervisor name ____________________________signature...........................................

Date of interview _________________________DD/MM/YY

Distance from your home to the health post ______________km

Distace from your home to the nearest health center ___________km

Distance from your home to nearest hospital ______________km

Distance from your home to the main road_____________Km

Result of the interview:

1. Completed

2. No woman at home

3. No all family at home

4. Another appointment

5. Refused

Age of child in complete months _____________or specify date of birth --------------------- (DD/MM/YY)

| Interviewer: I am going to start by asking you some questions about you and your household  [*Circle the answer among alternatives OR fill in the blank space]* | | | | | | | |
| --- | --- | --- | --- | --- | --- | --- | --- |
| *Household Characteristics* | | | | | | | |
| S.N | Question | | Response and Code | | | | Skip |
| 101 | How many family members are there in this household, including wife and husband? *Include only permanent residents(living greater than 6 months)* | | Family members in number . . . . . . . . … | | | |  |
| 102 | What is the main source of drinking water for members of your household? | | 1. Piped water 2. Dug well 3. Spring 4. River/stream   Others(*Specify*)____________ | | | |  |
| 103 | Where is that water source located? | | 1. In own compound 2. Elsewhere | | | | 105 |
| 104 | How long does it take to go there, get water, and come back? | | Minutes.................................  99. Don’ t know | | | |  |
| 105 | What do you usually do to make the water safer to drink?  RECORD ALL MENTIONED | | 1. Nothing 2. Boiling 3. Add bleach/Chlorine 4. Filter through cloth   Others (Specify)………….. | | | |  |
| 106 | What kind of toilet facility do members of your household usually use? | | 1. No facility but bush/open field 2. Flush toilet 3. pit latrine   Other(specify)_________ | | | |  |
| 107 | What type of fuel does your household mainly use for cooking?  [ MULTIPLE OPTION POSSIBLE] | | 1. Electricity 2. Bio gas 3. Kerosene 4. Charcoal 5. Wood 6. Animal dung   Others (Specify)……………… | | | |  |
| 108 | What type of fuel does your household mainly use for light sources?  [ MULTIPLE OPTIONS POSSIBLE] | | 1. Electricity 2. Biogas 3. Kerosene lamp 4. Solar light   Others (Specify)……………… | | | |  |
| 109 | The main material of the roof of the main house  Record OBSERVATION | | 1. Corrugated iron sheet 2. Thatch/leaf   Other(specify)_______ | | | |  |
| 110 | The main material of the floor of the main house is made of (observation)  *Write ONLY ONE answer* | | 1. Earth/ mud 2. ceramic tiles 3. cement   Other [specify]_________________ | | | |  |
| 111 | The main material of the walls (observation)  *Write ONLY ONE answer* | | 1. wooden and mud 2. stone with lime/cement/ bricks 3. Wood plank   Other [specify]________________ | | | |  |
| 112 | What is the primary source of income for this household?  *Circle ONLY ONE answer* | | 1. Farming, including livestock 2. employment/salary 3. petty trading (including the sale of fire-wood, charcoal, grass, etc) 4. Daily laborer   Other [specify]_________________ | | | |  |
| 113 | How many of the following animals does this household own?  IF NONE, RECORD '00'.  [PROBE AND MARK THAT ALL APPLY, MULTIPLE ANSWER IS POSSIBLE] | | Animal type | | | Amount |  |
|  |  |  | Cows/oxen /other cattle | | | _______ |  |
|  |  |  | horses/donkeys/mules | | | ___________ |  |
|  |  |  | Goats/Sheep | | | _________ |  |
|  |  |  | Chickens | | |  |  |
|  |  |  | Bee hives | | |  |  |
| 114 | Does any member of this household own any agricultural land? | | 0. No 1. Yes | | | | If No skip to Q. 116 |
| 115 | How many hectares/or “Timad” of agricultural land do members of this household own? ***If none, record '00'.*** | | Hectares____________ OR  “Timad”___________ | | | |  |
| 116 | Does any member of this household have an account with a bank/micro finance? | | 0. No | 1. Yes | | |  |
| 117 | Is the house listed as model farmer? | |  | |  | |  |
| 118 | Does your household have the following? | |  | |  | |  |
|  | 118.1 | Radio/ Television |  | |  | |  |
|  | 118.2 | Telephone; landline/ mobile |  | |  | |  |
|  | 118.3 | Bed with cotton/sponges/spring  Mattress |  | |  | |  |
|  | 118.4 | An animal-drawn cart/Bicycle/Motor  Bike / Bajaj/ car? |  | |  | |  |
|  | 118.5 | Sofa/ chair with Arm or backrest |  | |  | |  |

| **Section 2: Demographic characteristics of neonates** | | | |
| --- | --- | --- | --- |
| **No.** | **Questions and Filters** | **Coding Categories** | **SKIP** |
| 201. | What is the sex of your child? | 1. Female 2. Male |  |
| 202. | Where was the child born? | 1. Home  2. Health post  3. Health center  4. Hospital  Other :________________________ |  |
| 203. | How many were born in this birth? | 1. Singleton  2. Twins  3. Triple  4. Multiple |  |
| 204 | The outcome of this pregnancy? | 1. Miscarriage  2. Stillbirth  3. Live birth |  |
| 205. | Was (NAME) born early, late, or at the expected time? | 1. Early  2. On-time  3. Late |  |
| **Section 3:To determine the level of health care services received by neonates of mothers in the selected districts** **o****f Sidama Regionin2023.**   1. **I`m going to ask you the health care services receivedby neonate starting birthup** 2. **to end ofneonatal period** 3. | | | |
| 301. | Did yourinfant`s face wiped immediately after birth? | 0. No 1. Yes 99. Do not know |  |
| 302. | If yes, what was the materials used to wipe your infant`s face immediately after birth? | 1. Clean cloths 2. Cotton 3. Previously used cloth/towel 4. Others: _______________ |  |
| 303.* | Where did the newborn placed immediately after birth? | 1. On the mother’s chest or belly 2. On the newborn table 3. With someone else 4. Beside the mother 99. Don’t know   Others:__________________________________ |  |
| 304. | Did your newborn body dried immediately after delivery? | 0. No 1. Yes |  |
| 305.* | How was the newborn`sbody dried? | 1. Immediately and thoroughly after expulsion of neonate 2. Immediately after delivery of placenta 99. Do not know   Others:_______________________ |  |
| 306. | What was the materials used to dry your neonate`s body? | 1. New cloth/towel 2. Previously used but washed cloths 3. Previously used but not washed cloths   Others:__________________ |  |
| 307. | Did your neonate draped immediately after changing the wet cloth? | 0. No 1. Yes |  |
| 308. | If yes, what was the materials used to drape your neonate? | 1. New cloth/towel 2. Previously used but washed cloths 3. Previously used but not washed cloths   Others: __________________ |  |
| 309. | What was the instrument that used to cut the cord? | 1. New razor blade 2. Boiled razor blade 3. Previously used un boiled razor blade 99. Don`t know   Others:_____________________ |  |
| 310.* | When did your neonate`s cord clamped or tied? | 1. Immediately after delivery of the fetus 2. Immediately after delivery of placenta 3. Delayed for three minutes   99. Do not know  Others:______________________ |  |
| 311. | What was the materials used to clamp or tie your neonate`s cord? | 1. Not ties 2. String/ thread 3. Fiber from Ensete’ plant 99. Don’t now   Others:____________________ |  |
| 312.* | When did you start breast feeding your new born? | 1. Within one hour after birth 2. After one hour 3. At 24h after birth 4. At 48h after birth   99. Do not know  5. Others:___________ |  |
| 313. | Did your child weight checked immediately after birth? | 0. No 1. Yes |  |
| 314. | If yes, was the weight small or below 1.5kg? | 0. No 1. Yes |  |
| 315. | Was your child assessed for prematurity? | 0. No 1. Yes |  |
| 316. | In which of the following time or days did the child received neonatal health care?  Multiple choice can possible | Yes No  1. 6 hours after birth 1 0  2. 24 hours after birth 1 0  3. On day 3 after birth 1 0  4. Days between 7-14  after birth 1 0  5. Six weeks after birth 1 0 |  |
| 317. | Did your neonate checked for general physical examination? | 0. No 1. Yes |  |
| 318. | Did your child developed danger sign during the neonatal Period? | 0. No 1. Yes |  |
| 319. | Did your child had congenital anomaly? | 0. No 1. Yes |  |
| 320. | Did the newborn umbilical cord checked for bleeding? | 0. No 1. Yes |  |
| 321. | What substance did you applied to your newborn`s cord? | 1. Butter applied 2. Vaseline 3. Nothing applied 4. Other:_______________________ |  |
| 322. | How did you handled the umbilical cord after cut? | 1. Without dressing 2. With covering 3. Other: ____________________ |  |
| 323. | What did you feed your newborn baby first three days? | 1. Breast milk/colostrums 2. Plain water 3. Sugar water 4. Fresh butter 99. Don’t know 5. Other;________________ |  |
| 324. | In what pattern did you breast feed your newborn? | 1. On demand 2. Not on demand 99. Do not know 3. Other:__________ |  |
| 325. | Did you continued exclusive breast feeding? | 0. No 1. Yes |  |
| 326. | If No, for Q.325. What were your reasons? | 1. Health professional’s recommendation 2. Feeling of insufficient amount of breast milk 3. Maternal illness or 4. Use of medication 5. Infant illness 6. Return to work 7. Other:________________________ |  |
| 327. | If yes, for Q.325, for how long did you exclusively feed your breast milk? | 1.For 03 months  2. For 04 months  3. For 05 months  4. For 06 months  99. Do not know  5. Others: __________________ |  |
| 328. | If Yes, for Q.No.325. How frequently feed your breast milk?  Probe: in how many hours interval | 1. Every two to three hours 2. When the baby wants 3. When the baby cry’s 4. When the baby awake from sleep   Other:__________ |  |
| 329. | Did your baby received health care from the trained person within 24hrs? | 0. No  1. Yes | If No, skip to Q 330 |
| 330. | If yes for Q.328, what did the trained persons to your baby? | 1.Performed general physical examination  2.Given immunization  3. Checked umbilical cord bleeding  4. Done temperature measurement  5. Checked breastfeeding difficulties  Others: _________________ |  |
| 331. | When did you washed your newborn after birth? | 1. After 24hour 2. Within one hour following delivery 3. After 2 hours following delivery 99. Don’t know   Others:________________ |  |
| 332. | Did your child`s got fever when you touch during neonatal period? | 0. No 1. Yes |  |
| 333. | When did you started sun light exposure to your new born? | 1.After two weeks  2.After 3 weeks  3.After a month  99.Do not know  Others:_________________ |  |
| 334. | At what time your newborn received OPV immunization | 1. At birth 2. At 45 days 3. After 45 days 4. At 10^th^ week   Others:_______________________ |  |
| 335. | At what time your newborn received BCG immunization? | 1. At birth 2. At 45 days 3. After 45 days 4. At 10^th^ week   Others:_______________________ |  |
| 336. | How did you keep clean your newborn eye? | 1. Clean the eye separately with clean cloth 2. Clean with finger 3. Clean with clean close together with other parts of the face 99. Don’t know   Others:___________________ |  |
| **Section - 4.To assess neonatal illnesses identified, managed and referred in the community of selected districts in Sidama region in 2023.** | | | |
| 401. | Did your neonate started breathing/crying on his/her own immediately after delivery? | 0. No  1. Yes | If yes, skip to Q.no. 407 |
| 402. | If No for Q. 401, who ensured baby was not breathing? | 1.Health extension worker  2. Midwife  3. Nurse  4. Doctor  Other:_______________ |  |
| 403. | If health extension worker, what action did they took? | 1.Referral to hospital  2.Managed by their own  Other :__________________ |  |
| 404. | Did your baby receive medical care from other health professional in health facility? | 0. No  1. Yes | If No skip to Q.406 |
| 405. | If Yes, who provided the medical care? | 1. Health extension worker 2. Midwife 3. Nurse 4. Doctor   Other:____________ |  |
| 406. | What was the outcome of the newborn? | 1.Improvement  2.Sickness  3. Death |  |
| 407. | Did the health extension worker measure your newborn`s body temperature during the neonatal period? | 0. No  1. Yes | If No skip to Q. 410. |
| 408. | Was the newborn`s body temperature normal? | 0. No 1. Yes | If Yes, skip to Q.410 |
| 409. | If No for Q.408, what action did health extension worker take? | 1. The health extension worker managed it.  2. The health extension referred to hospital  3. Nothing done  Others:___________________ |  |
| 410. | Did health extension worker measure baby`s weight? | 0. No  1. Yes | If No. skip to Q. 413 |
| 411. | If yes for Q. 410, was the weight of the newborn normal as of previously born children? | 0. No 1. Yes | If yes, skip to Q.413 |
| 412. | If No for Q.411, what was the action took by health extension worker? | 1. The health extension worker managed  2. The health extension referred to hospital  3. Nothing done  Others:___________________ |  |
| 413. | After the birth of your baby, did health extension worker done physical examination? | 0. No  1. Yes | If No, skip to Q.417 |
| 414. | Did your newborn had one of the following characteristics after birth?  Multiple answers possible | - Yes No - 1.Thin, shiny, pink or red skin 1 0 - 2. Little body fat 1 0 - 3. Little scalp hair 1 0 - 4. lots of lanugo (soft body hair) 1 0 - 5. Weak cry and body tone 1 0 - 6. Small and underdeveloped genitalia 1 0 - 7. No any of the above conditions identified - Other:__________________________ |  |
| 415. | How did the health extension manage it? | 1. Managed using KMC  2. Referred to hospital  3. Nothing done  Others:___________________ |  |
| 416. | How long after the delivery was this done? | \|__\|__\| Minutes  \|__\|__\| Hours  \|__\|__\| Days  \|__\|__\| Weeks  \|__\|__\| Months |  |
| 417. | Did your child got illness/problem during neonatal period? | 0. No 1. Yes | If No, skip to Q. 420. |
| 418. | Which of the following illnesses/problems identified? | No Yes   1. Neonatal sepsis 0 1 2. Pneumonia 0 1 3. Birth asphyxia 0 1 4. Hypothermia 0 1 5. Preterm 0 1 6. Low birth neonates 0 1   Birth defects: |  |
| 419. | What action did the health extension workers took? | No Yes   1. Appointment 0 1 2. Referral 0 1 3. Management 0 1 4. Follow-up 0 1   Others:____________ |  |
| 420. | Was there any identified birth defect/s? | 0. No 1. Yes | If No. skip to Q. 423 |
| 421. | If yes for Q.420, which birth defect/s was/were identified? | 1. Anencephaly 2. Spinal bifida 3. Cleft lip 4. Cleft palate 5. Anorectal atresia/stenosis   Others:_______________ |  |
| 422. | What action did the health extension workers took? | No Yes   1. Appointment0 1 2. Referral 0 1 3. Management0 1 4. Follow-up0 1   Others:____________ |  |
| 423. | Did your child develop any danger sign/s during neonatal period? | 0. No 1. Yes | If No, skip to Q. 428 |
| 424. | Which of the following danger sign/s did your child developed during the neonatal period?  Multiple answer is possible | No Yes  1. Not sucking properly 0 1  2. Convulsion 0 1  3. Fast breathing  (breathing rate 60 per minute) 0 1  4.Difficult to wake up, no  spontaneous movement 0 1  5. Red cord stump or with pus 0 1  6. Severe chest in-drawing 0 1  7. Fever (temperature 37.5 C) 0 1  8. Low body temperature  (temperature <35.5 °C) 0 1  9. Red/ discharging eye 0 1  10. Yellowish eye, skin, palms  and soles in first 24 hours of  life 0 1  11. Lethargy 0 1  12. Diarrhea 0 1  13. Persistent vomiting 0 1  Other:________________ |  |
| 425. | If yes for Q no.423, where did you go to seek care? | 1.Health post  2.Health center  3.Hospital  Other:________________________ | If not health post skip to Q. 428 |
| 426. | If yes for Q no.423, who did the assessment? | 1.Health extension worker  2. Midwife  2. Nurses  3. Health officer  4. Doctor  99. Do not know  Others:______________________ |  |
| 427. | If the health extension worker did the assessment in the Q. no.426, what action did she took? | 1 Treat with antibiotics  2. Referral  99. Do not know  Others:__________________________ |  |
| 428. | Did your child have cough during the neonatal period? | 0. No  1. Yes | If no, stop the interview here. |
| 429. | If yes, would you tell us the duration of the illness? | _________days |  |
| 430. | Did he/she had an unusual difficulty of breathing or fast breathing? | 0. No  1. Yes |  |
| 431. | Who did the examination? | 1.Health extension worker  2.Traditional birth attendants  3. Nurses  4. Midwifes  5. Doctor  Others:___________________________ | If HEWs is not examined stop the interview here. |
| 432. | If the health extension workers examined in the Q.431, What was the action taken? | 1.Referral  2. Treating with antibiotics  3. Appointment  99. Do not know  Other:______________________ |  |
